# Observed and self-reported COVID-19 health protection behaviours on a university campus and the impact of a single simple intervention

**DOI:** 10.1101/2021.06.15.21258920

**Authors:** Rachel Davies, John Weinman, G James Rubin

**Author notes:** Competing interests: None declared.

## Abstract

**Objectives:** Hygiene behaviours had been an essential component of attempts to slow the spread of SARS-CoV-2. Most data on adherence to these behaviours is collected via self-reporting, which can differ from observed behaviours. We quantified this discrepancy among a university sample and tested the impact of simple intervention on observed behaviour.

**Study design:** Cross-sectional questionnaire of behaviour on campus compared to direct observation on one day without, and one day with, additional signage.

**Methods:** We circulated an email inviting all staff and students at our university to complete a questionnaire asking how often they wear a mark or practice hand hygiene when entering campus buildings, and how often they practiced social distancing within campus buildings. We observed all entrants to the main building on one campus on a baseline day and on a day after installing a large sign reminding people that these behaviours were mandatory.

**Results:** In our survey, 172 out of 252 respondents (68%) reported always cleaning their hands, 225 out of 251 (90%) reported always wearing a face covering, and 124 out of 252 (49%) reported always maintaining social distancing. On the baseline day of observation, 50 out of 311 people (16.1%) cleaned their hands and 256 (82.3%) wore a face covering correctly. Out of 119 people whom we could assess, 9 (7%) maintained social distance from others. The signage was associated with significant improvements for cleaning hands (104 / 375 people: 27.7%), wearing a face covering (374 / 375; 99.8%) and maintaining social distance (79 / 144; 54.8%).

**Conclusions:** Greater use of observational methods will provide a more accurate measure of behaviour than the current reliance on self-report and allow interventions to be robustly tested.

---

The promotion of hygiene and behavioural adaptations among the general public has been an essential component of attempts to mitigate the impact of the COVID-19 pandemic^1,2^. Attempts to understand adherence with various recommended behaviours, and factors associated with adherence, have relied on the use of cross-sectional surveys, often commissioned via market research organisations^3^. While these surveys can provide useful insight into trends over time, the accuracy of their data is often unclear, particularly for behaviours that may occur many times a day, in many different settings, and are therefore easy to misremember^4,5^. In this study, we compared data from a cross-sectional survey of students at one large university in London with data based on direct observation of the same behaviours. We also tested whether a simple intervention (the installation of clear signage) could improve adherence to these behaviours. The specific behaviours we explored were all mandatory at the university: cleaning hands upon entering a building; wearing a face covering upon entering a building; and maintaining social distancing.

We used two study designs. First, we used an online cross-sectional survey to measure self-reported adherence to our three behaviours. Second, we used direct observation of behaviour at the entrance to a campus building on two days. Day one consisted of a control condition. One day two we installed a large sign at the entrance reminding people about the university rules. This study was approved by King’s College London’s BDM Research Ethics Subcommittee.

For the survey, an invitation to participate was disseminated via a biweekly newsletter advertising research studies within the university. This is sent to all students and staff. The survey consisted of 16 items. Among other things, we asked:

- How often do you wear a mask when entering the [university] campus buildings?
- How often do you practice hand hygiene measures when entering the [university] campus buildings?
- How often do you practice social distancing guidelines when within the [university] campus buildings?

Response options consisted of ‘always’, ‘often’ ‘sometimes’ ‘rarely’ and ‘never’. Responses were obtained between 1 December 2020 and 22 March 2021.

For the observational study, one of us based ourselves outside the sole entrance to the main campus building on two consecutive days. For day two we erected a sign immediately outside the entrance stating the mandatory policy for mask wearing, hand-hygiene and social distancing within the building. (Supplementary figure 1 and 2). Except for a short break for lunch, every person who entered the building between 9am and 5pm was observed. Participants were observed for 3 behaviors as they entered the building: hand hygiene, adequate face covering wearing and adequate social distancing. Hand hygiene was defined as use of hand sanitizer or gel or use of a hand washing station. The observer was able to see if the person used their own supply, or if they used a hand sanitizer dispenser or a recently installed sink that were available immediately inside the entrance. Adequate wearing of a face covering was defined as the wearing of any covering that covered both the mouth and nose of the participant. Adequate social distancing was defined as remaining >2m away from others within the building. In some instances, social distancing of participants could not be assessed, for example if an individual entered the building alone, and the entrance area was empty of others. If groups entered together, and were not socially distanced before entering the building and on approach, they were assumed to be a bubble/household. No identifiable or personal information was collected and participants remained anonymous throughout.

In total, 252 participants responded to the cross-sectional survey, 311 people were observed on day one and 375 people were observed on day two. Table one shows the number reporting each of the three behaviours in the survey together with the number of people performing each behaviour on days one and two of the observational study. Comparing self-reported behaviours with behaviour observed on day one of our experiment, it can be seen that many more people reported “always” practicing hand hygiene (68%) or socially distancing (49%) than was actually observed (16% and 7%, respectively), but that self-reported rates of face covering wearing were more accurate (90% vs 82%). Adherence for all behaviours was significantly better on day two of the experiment, after our sign was in place.

**Table one:**
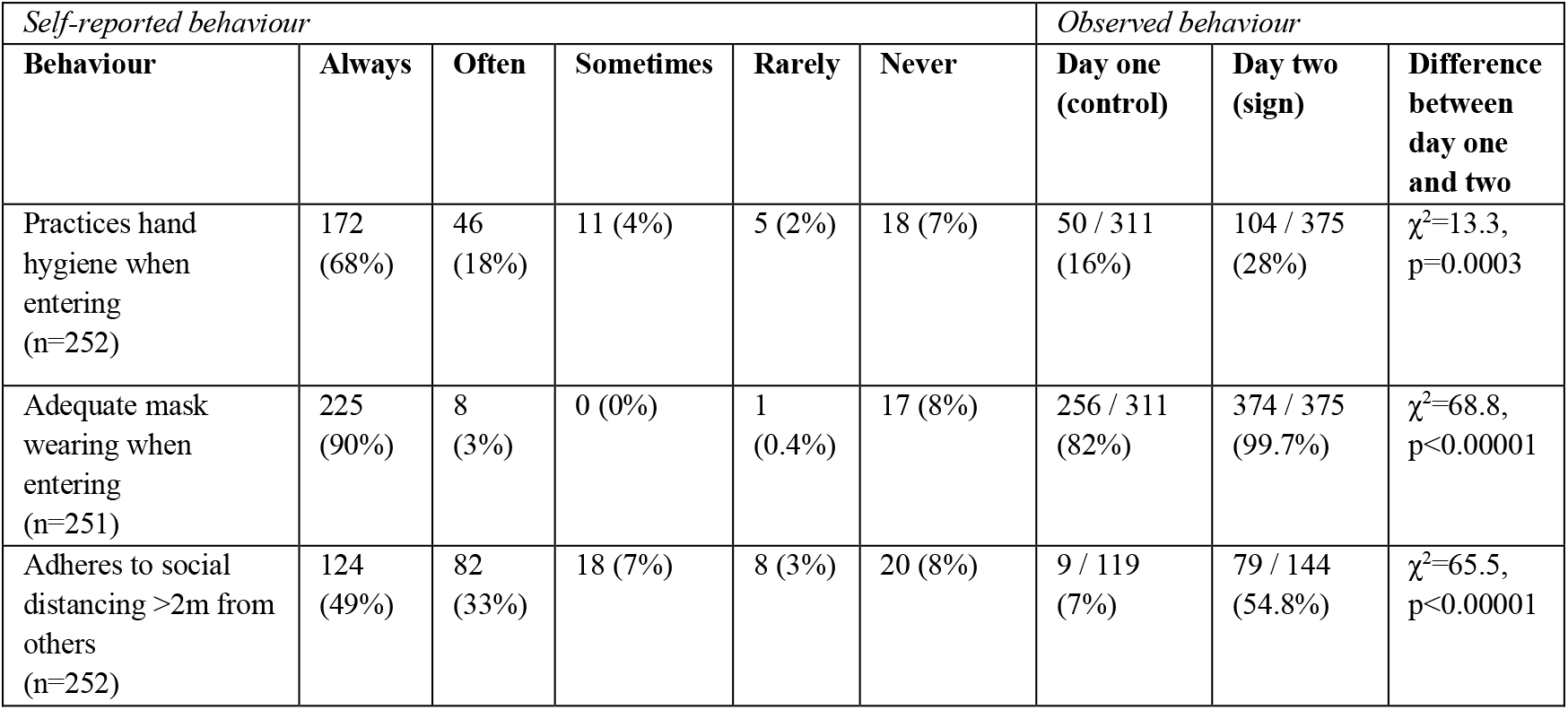
Behaviour data

| *Self-reported behaviour* |  |  |  |  |  | *Observed behaviour* |  |  |
| --- | --- | --- | --- | --- | --- | --- | --- | --- |
| **Behaviour** | **Always** | **Often** | **Sometimes** | **Rarely** | **Never** | **Day one**<br>**(control)** | **Day two**<br>**(sign)** | **Difference**<br>**between**<br>**day one**<br>**and two** |
| Practices hand hygiene when entering (n=252) | 172 (68%) | 46 (18%) | 11 (4%) | 5 (2%) | 18 (7%) | 50 / 311 (16%) | 104 / 375 (28%) | $\chi^2=13.3$,<br>p=0.0003 |
| Adequate mask wearing when entering (n=251) | 225 (90%) | 8 (3%) | 0 (0%) | 1 (0.4%) | 17 (8%) | 256 / 311 (82%) | 374 / 375 (99.7%) | $\chi^2=68.8$,<br>p<0.00001 |
| Adheres to social distancing >2m from others (n=252) | 124 (49%) | 82 (33%) | 18 (7%) | 8 (3%) | 20 (8%) | 9 / 119 (7%) | 79 / 144 (54.8%) | $\chi^2=65.5$,<br>p<0.00001 |

Our data demonstrate that self-report surveys can substantially overestimate adherence to hygiene recommendations. This was particularly the case for hand hygiene and social distancing. Only rates of observed face covering wearing matched the participants’ self-reports. We speculate that this is because, as a clearly visible action, it is more likely to be enforced by campus security and / or noticed by others, leading to higher adherence.

Our study also demonstrates that observational methods can be readily used to evaluate the impact of simple interventions to improve adherence, providing a better indication of where problems lie and what interventions work to resolve them. This is consistent with other evidence on the efficacy of the use of notices and other environmental prompts for activating behaviour changes, including hand hygiene^6^.

There are several limitations to this work. The self-report study was not necessarily representative of larger scale, national surveys conducted in this field, particularly as we did not use quota sampling or weighting. The observational study was limited by its focus on a single entrance to one university building. Other populations or other buildings may have higher adherence. Difficulties assessing whether people were in household bubbles made our assessment of social distancing particularly challenging, and results based on these data in particular should be taken as tentative.

## Data Availability

N/A

