## Supplementary figures and images for "Observed and self-reported COVID-19 health protection behaviours on a university campus and the impact of a single simple intervention"

### supplementary figure 1.png

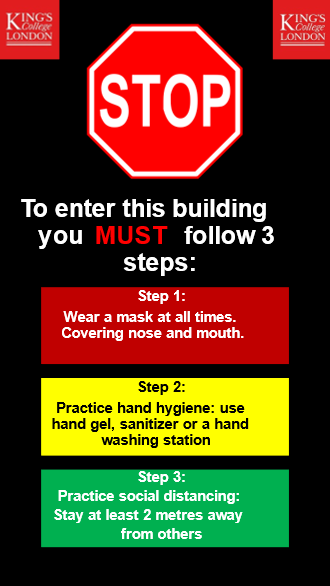

### supplementary figure 2.jpg

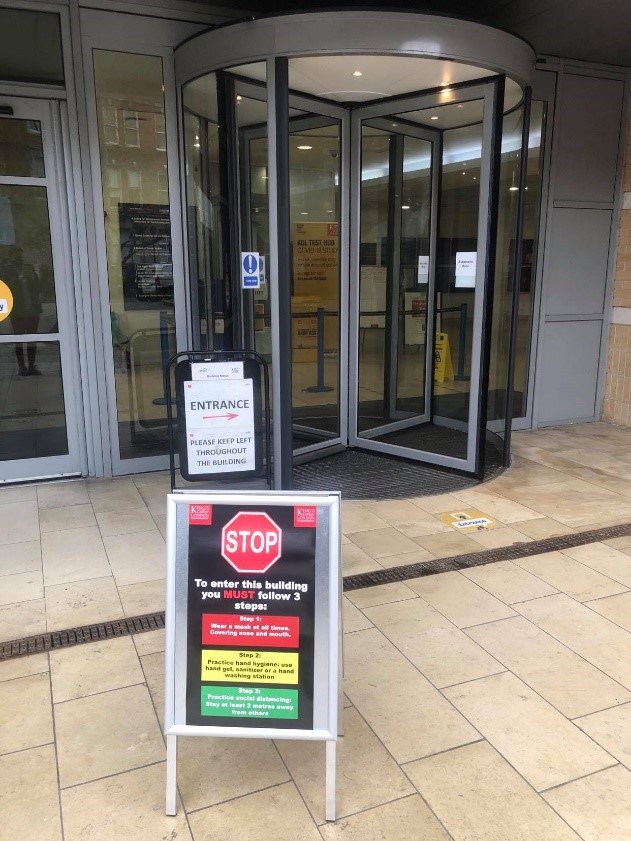
